# The Clinical-Functional Vulnerability Index-20 (IVCF-20) Predicts Adverse Outcomes in Older Adults Admitted through the Emergency Department

**DOI:** 10.1101/2025.11.05.25339627

**Authors:** Tatiana C. E. Pinheiro, Marco A. F. Angelo, Márlon J. R. Aliberti, Carla J. Machado, Vicente E. Gonçalves, Nádia E. Ramos, Camila O. Alcântara, Fabiano M. Pereira, Ana P. A. F. Peixoto, Fátima S. Machado, Bernardo M. Viana, Edgar N. Moraes, Maria A. C. Bicalho

**Affiliations:** Reference Center for Older Adults, Hospital das Clínicas, Universidade Federal de Minas Gerais (UFMG), Belo Horizonte, Minas Gerais, Brazil; Faculdade de Medicina, Universidade Federal de Minas Gerais (UFMG), Belo Horizonte, Brazil; Hospital Risoleta Tolentino Neves, Universidade Federal de Minas Gerais (UFMG), Belo Horizonte, Brazil; Secretaria de Estado da Saúde de Minas Gerais, Brazil; Laboratório de Investigação Médica Em Envelhecimento (LIM-66), Serviço de Geriatria, Hospital das Clínicas (HCFMUSP), Faculdade de Medicina, Universidade de São Paulo (USP), São Paulo, Brazil; Research Institute, Hospital Sírio-Libanês, São Paulo, Brazil; Instituto Neurotecnologia Responsável (INCT-Neurotec R), Conselho Nacional de Desenvolvimento Científico e Tecnológico (CNPq), Brazil

**Keywords:** frailty, Emergency Department, screening, prognosis, risk score

## Abstract

**Background:** Frailty assessment in the Emergency Department (ED) is essential for identifying older adults at risk of adverse outcomes. The 20-item Clinical-Functional Vulnerability Index (IVCF-20) is a rapid, multidimensional screening tool widely used in Brazilian primary care, but its predictive validity in the ED has not been established. We aimed to evaluate the ability of the IVCF-20 to predict 180-day mortality and other adverse outcomes in older adults admitted to a public ED.

**Methods:** Observational cohort study comprising patients aged ≥60 years consecutively admitted through the ED of a University Hospital in Brazil. Baseline frailty was assessed with the IVCF-20 and categorized as robust (0-6), pre-frail (7–14), mild-to-moderate (15–29), and severe frail (30–40). The Clinical Frailty Scale (CFS), a validated frailty tool in the ED, was also applied. The primary outcome was 180-day mortality; secondary outcomes included in-hospital and 90-day mortality, prolonged length of stay, home care referral, and ED revisit or hospital readmission. Logistic regression estimated associations between frailty and 180-day mortality, Kaplan-Meier curves illustrated survival across frailty levels. ROC analyses evaluated secondary outcomes.

**Results:** A total of 310 patients with a median age of 72 years, 58.1% were male. Frailty prevalence ranged from 53.9% (IVCF-20) to 60.1% (CFS). The IVCF-20 score was independently associated with 180-day mortality (adjusted OR = 1.06; 95% CI = 1.02–1.10; p = 0.002). Severely frail participants had an 8.4-fold higher risk of death than robust individuals (adjusted OR = 8.37; 95% CI = 2.20–31.81). Kaplan-Meier curves showed a graded mortality increase across IVCF-20 categories. Both instruments predicted secondary outcomes, though CFS demonstrated slightly better discrimination for mortality.

**Conclusions:** IVCF-20 predicted 180-day mortality, home care referral, and ED revisit/readmission. Its rapid, judgment-free format supports its feasibility for frailty screening at the ED.

## Introduction

The increasing number of older adults admitted to the Emergency Department (ED) is a global phenomenon, occurring faster than overall population growth ^1^. In Brazil, the mean age of ED patients has progressively increased, and older adults already account for more than 26% of admissions ^2,3^. Compared with younger adults, they present a higher risk of adverse outcomes, including prolonged length of stay (LOS), functional decline, readmission, and death ^4^. However, there is substantial heterogeneity in physiological reserve among older adults, influencing treatment response and prognosis ^5^. Identifying older adults at higher risk is essential to guide clinical decisions, allocate resources efficiently, and prevent iatrogenesis ^6^.

Acute deterioration criteria are often used to predict outcomes in older adults admitted to the ED ^7^. Nevertheless, early warning scores may be less reliable in this population, who frequently present with atypical clinical manifestations ^7,8^. Traditional predictors, such as advanced age and comorbidities, are insufficient to adequately determine clinical-functional vulnerability ^9,10^. Baseline frailty provides a more accurate measure of physiological reserve than chronological age, offering greater insight into individual prognosis and supporting care planning ^11–13^.

Frailty, defined as a progressive decline across multiple health domains and diminished resistance to stressors ^14^, increases vulnerability to repeated admissions, functional dependence, delirium, falls, fractures, and death ^5,15^. The Comprehensive Geriatric Assessment (CGA) is the gold standard for identifying frailty in community settings, but its applicability in the ED is limited by the time constraints and the need for specialized teams ^16^. In high-flow emergency settings, with high bed demand and prolonged waiting times for admission ^17^, patients may also be unable to complete physical or performance-based tests ^18^.

Frailty screening instruments have been studied regarding their applicability and feasibility in the ED ^19^. The 20-item Clinical-Functional Vulnerability Index (IVCF-20) is a rapid, multidimensional frailty screening tool that correlates strongly with the CGA. Validated in primary health care settings, the instrument has shown excellent performance in identifying frail older adults ^20^. It is among the most frequently analyzed tools for frailty detection based on its clinimetric properties ^21^ and has been adopted by the Brazilian Ministry of Health as the official national tool for classifying frailty among older adults ^22^. However, its predictive validity for adverse outcomes has not been previously investigated. The aim of this study is to assess the ability of the IVCF-20 to predict mortality and other adverse outcomes among older adults admitted to a public ED.

## Methods

### Study design and Setting

We conducted a longitudinal observational study including two independent prospective cohorts of participants aged ³ 60 years who were admitted to the ED for clinical or surgical emergencies and remained at least one night in this public university hospital unit. The first cohort was followed from December 2019 to May 2020 and from November 2020 to January 2021, as previously described ^13^; and, the second cohort was followed from July to December 2023 and is part of the multicenter “Creating a Hospital Assessment Network in Geriatrics” (CHANGE) Study ^23^.

The University Hospital of the Federal University of Minas Gerais (UH-UFMG) is a 460-bed public hospital and referral center for medium- and high-complexity care in Minas Gerais, Brazil. The ED provides care for patients with clinical and non-trauma-related surgical emergencies referred from the specialized outpatient clinics of the UH-UFMG complex, and from urgent care units in Belo Horizonte, Minas Gerais, Brazil.

The first cohort Study was approved by the Research Ethics Committee of the Federal University of Minas Gerais (UFMG), under the protocol 23649519.0.0000.5149. The CHANGE Study was approved by the Research Ethics Committee of the University of São Paulo (USP), under the protocol 56802322.9.1001.0068.

### Participants

During the study periods, the ED recorded a monthly average of approximately 926 visits, of which 37.4% were older adults. A total of 749 individuals aged ³ 60 years who were admitted to the ED and remained in the hospital for at least one night met the eligibility criteria. We excluded patients who were unable to communicate (those who were actively dying or had advanced dementia without a caregiver able to provide consent and reliable information) (n=40), those transferred to other units, discharged, or deceased within 24 hours of admission (n=340), individuals with terminal illness and life expectancy < 6 months in the second cohort (n=34), and those who declined participation (n=25) (Supplementary Figure S1). Written informed consent was obtained from all participants or from their legal representatives in cases of cognitive impairment.

Follow-up data were collected 3 to 6 months after hospital discharge through telephone interviews conducted by the researcher team, complemented by review of medical records. At the end of the 6-month follow-up, mortality status was verified for all participants.

### Variables and Measurements

Sociodemographic data, including age, sex, race, education level, and family arrangement, were collected for analysis. The severity of the acute illness was assessed using the National Early Warning Score 2 (NEWS2) ^24^. Comorbidities were recorded according to the Charlson Comorbidity Index (CCI) ^25^. Baseline frailty was defined as the patient’s state of vitality or frailty relative to two weeks before hospital admission or before the onset of the acute illness leading to hospitalization ^26^. Frailty was assessed using two instruments: the IVCF-20 ^20,27^ (Supplementary Figure S2) and the Clinical Frailty Scale (CFS) ^28,29^.

For the IVCF-20, a cutoff score of ≥15 was considered indicative of frailty, as it is the most useful for identifying frailty, with 52% sensitivity and 98% specificity ^27^. Participants were categorized according to the original IVCF-20 scores as robust (0–6), pre-frail (7–14), and frail (15–40). To explore prognostic differences among frail individuals, the frail category was subdivided in mild-to-moderate (15–29) and severe (30–40) frailty. Physical measurements, including weight, height, and gait speed, were not collected for all participants because these assessments may not be feasible in acutely ill patients and in high-flow emergency settings ^18^. Therefore, item 14 (aerobic capacity) was instead assessed by self-reported weight loss and by calf circumference. The latter measure was considered more feasible, requiring only a simple measuring tape - widely available, quick to perform, and applicable even for bedridden patients.

The CFS was also used as a rapid frailty assessment tool for predicting adverse outcomes in older adults in emergency settings. This widely adopted scale demonstrated good predictive ability for mortality and other adverse outcomes in older adults admitted due to clinical or surgical emergencies ^12,13,18^. Participants scoring 5–8 were classified as frail, and those scoring 9 were considered terminally ill, with a life expectancy of less than 6 months ^28,29^.

Data were obtained from patients and/or caregivers and collected by trained investigators using a standardized electronic form on the Redcap® platform ^30^.

### Outcome measures

The primary outcome was 180-day mortality. Secondary outcomes included in-hospital mortality, 90-day mortality, prolonged length of stay (LOS), referral to home care at discharge, institutionalization, and ED revisit or hospital readmission. Prolonged LOS was defined as the upper tertile of hospital LOS distribution in the study sample. ED revisit and hospital readmission were defined as unplanned returns to the ED or unplanned hospital readmissions within three months after discharge, respectively.

### Statistical Analysis

For all numerical variables, the Shapiro-Wilk test rejected the null hypothesis of normality. Continuous variables were expressed as medians and interquartile ranges, and categorical variables as absolute and relative frequencies. The Kruskal-Wallis test was used to compare continuous variables across IVCF-20 groups, and the chi-square or Fisher’s exact test (when appropriate) was applied for categorical variables.

The 180-day mortality outcome was analyzed as a dichotomous variable using binary logistic regression. Each independent variable was tested in univariate logistic regression models to predict mortality. A multivariate model was constructed, treating the IVCF-20 as a continuous variable. The instrument proved to be a linear continuous predictor in the logit of 180-day mortality, showing a progressive increase in mortality risk with higher scores. A second model treated the IVCF-20 as an ordinal variable to demonstrate the gradient of risk across frailty categories and to facilitate clinical interpretation. Finally, a third model was constructed using CFS as a continuous variable for comparison.

All models were adjusted for demographic and clinically relevant confounders with *P* < .20 in univariate analysis. Variables with *P* < .05 were retained in the final models. Age was not included in IVCF-20 models to avoid duplication of adjustment, as it is already incorporated into the instrument. Measures of association were expressed as odds ratio (ORs) for univariate analysis and adjusted OR (aORs) for multivariate analysis, with 95% confidence intervals (CIs). Model performance was compared using the area under the receiver operating characteristic (ROC) curve (AUC) and the DeLong test. Kaplan-Meier survival curves were generated to illustrate survival estimates across IVCF-20 categories. For a secondary analysis, we constructed a Cox proportional hazard model with IVCF-20 as a continuous variable; however, the proportional hazards assumption was not met.

For secondary outcomes, AUCs of both instruments were compared using DeLong test, limited to outcomes significantly associated in univariate analysis.

All analyses were performed using Jamovi® software (version 2.7.6.0, The Jamovi Project, Sydney, Australia) for Mac.

Data reporting is consistent with Strengthening the Reporting of Observational Studies in Epidemiology (STROBE) guidelines ^31^.

## Results

Our final sample reached 310 participants. The median age of participants was 72 years (IQR, 66–78); 58.1% were male, 62.6% self-identified as Black or Brown, and 75.2% had less than 8 years of education (Table 1). The main reasons for admission were dyspnea (18.7%) and acute neurological deficit (9.4%).

**Table 1.** Characteristics of patients and outcomes according to frailty status measured by the IVCF-20 (n = 310). Belo Horizonte, Minas Gerais, 2019–2021 and 2023–2024.

| Variables | Robust<br>(0-6)<br>n=45 | Pre-frail<br>(7-14)<br>n=98 | Mild-to-moderate<br>frail<br>(15-29)<br>n=143 | Severe<br>frail<br>(30-40)<br>n=24 | Total<br>n=310 | P -<br>value |
| --- | --- | --- | --- | --- | --- | --- |
| **Sociodemographic Characteristics** |  |  |  |  |  |  |
| Age, median<br>(IQR) | 70 (7) | 71 (7.75) | 72 (12.5) | 82 (10.25) | 72 (12) | < .001 |
| Sex, female, n (%) | 10 (22.2)<sup>b</sup> | 41 (41.8) | 65 (45.5) | 14 (58.3) | 130 (41.9) | .014 |
| Race, n (%) |  |  |  |  |  | .381 |
| White | 12 (26.7) | 31 (31.6) | 55 (38.5) | 12 (50.0) | 110 (35.5) |  |
| Brown | 26 (57.8) | 51 (52.0) | 58 (40.6) | 7 (29.2) | 142 (45.8) |  |
| Black | 6 (13.3) | 14 (14.3) | 27 (18.9) | 5 (20.8) | 52 (16.8) |  |
| Indigenous | 1 (2.2) | 2 (2.0) | 3 (2.1) | 0 (0.0) | 6 (1.9) |  |
| Education, n (%) |  |  |  |  |  | .932 |
| Illiterate | 5 (11.1) | 11 (11.2) | 23 (16.1) | 4 (16.7) | 43 (13.9) |  |
| 1–7 years | 28 (62.2) | 58 (59.2) | 89 (62.2) | 15 (62.5) | 190 (61.3) |  |
| 8–11 years | 10 (22.2) | 25 (25.5) | 26 (18.2) | 4 (16.7) | 65 (21.0) |  |
| 12 years or more | 2 (4.4) | 4 (4.1) | 5 (3.5) | 1 (4.2) | 12 (3.9) |  |
| Family arrangement, n (%) |  |  |  |  |  | < .001 |
| Lives with family | 37 (82.2) | 79 (80.6)<sup>b</sup> | 132<br>(92.3)<sup>a</sup> | 21 (87.5) | 269 (86.8) |  |
| Lives alone | 8 (17.8) | 19 (19.4)<sup>a</sup> | 11 (7.7)<sup>b</sup> | 0 (0.0) | 38 (12.3) |  |
| Institutionalized | 0 (0.0) | 0 (0.0) | 0 (0.0) | 3 (100)<sup>a</sup> | 3 (1.0) |  |
| **Clinical-Functional Characteristics** |  |  |  |  |  |  |
| CCI, median<br>(IQR) | 5 (2) | 6 (4) | 6 (3) | 9 (5) | 6 (3) | < .001 |
| DM with end-organ damage | 2 (4.4) | 14 (14.3) | 28 (19.6) | 4 (16.7) | 48 (15.5) | .104 |
| History of MI | 4 (8.9)<sup>b</sup> | 21 (21.4) | 30 (21.0) | 10 (41.7)<sup>a</sup> | 65 (21.0) | .017 |
| Heart failure | 8 (17.8)<sup>b</sup> | 32 (32.7) | 59 (41.3)<sup>a</sup> | 9 (37.5) | 108 (34.8) | .034 |
| Previous stroke or<br>TIA | 9 (20.0) | 12 (12.2) | 24 (16.8) | 7 (29.2) | 52 (16.8) | .220 |
| Metastatic cancer | 3 (6.7) | 14 (14.3) | 25 (17.5) | 4 (16.7) | 46 (14.8) | .333 |
| Dementia | 0 (0)<sup>b</sup> | 2 (2.0)<sup>b</sup> | 14 (9.8) | 14 (58.3)<sup>a</sup> | 30 (9.7) | < .001 |
| Delirium | 0 (0)<sup>b</sup> | 2 (2.0)<sup>b</sup> | 14 (9.8) | 8 (33.3)<sup>a</sup> | 24 (7.7) | < .001 |
| COPD | 5 (11.1) | 14 (14.3) | 25 (17.5) | 7 (29.2) | 51 (16.5) | .240 |
| Chronic liver<br>disease | 4 (8.9) | 4 (4.1) | 16 (11.2) | 3 (8.7) | 27 (8.7) | .183 |
| CKD (moderate to<br>severe) | 7 (15.6) | 25 (25.5) | 42 (29.4) | 7 (29.2) | 81 (26.1) | .318 |
| IVCF-20, median<br>(IQR) | 5 (3) | 11 (3) | 19 (7.5) | 31 (1.0) | 15 (12) | < .001 |
| CFS, median<br>(IQR) | 3 (1) | 4 (1) | 5 (1) | 7 (1.25) | 5 (2) | < .001 |
| Needs help with bathing | 0 (0.0)<sup>b</sup> | 0 (0.0)<sup>b</sup> | 34 (23.8)<sup>a</sup> | 23 (95.8)<sup>a</sup> | 57 (18.4) | < .001 |
| Hospitalization in past 6 months | 8 (17.8)<sup>b</sup> | 49 (50.0) | 93 (65.0)<sup>a</sup> | 19 (79.2)<sup>a</sup> | 169 (54.5) | < .001 |
| Polypharmacy | 13 (28.9)<sup>b</sup> | 60 (61.2) | 110 (76.9)<sup>a</sup> | 23 (95.8)<sup>a</sup> | 206 (66.5) | < .001 |
| ≥2 falls in past year | 0 (0)<sup>b</sup> | 13 (13.3)<sup>b</sup> | 45 (31.5)<sup>a</sup> | 13 (54.2)<sup>a</sup> | 71 (22.9) | < .001 |
| ≥5 chronic diseases | 6 (13.3)<sup>b</sup> | 33 (33.7) | 63 (44.1)<sup>a</sup> | 16 (66.7)<sup>a</sup> | 118 (38.1) | < .001 |
| Depressed mood/ Anhedonia | 3 (6.7)<sup>b</sup> | 25 (25.5)<sup>b</sup> | 90 (62.9)<sup>a</sup> | 16 (66.7) | 134 (43.2) | < .001 |
| NEWS2, median (IQR) | 1 (2) | 2 (2) | 3 (5) | 5 (3) | 2.5 (4) | < .001 |
| Length of stay, median (IQR) | 9 (15) | 8 (12.75) | 9 (14) | 9 (12.25) | 9 (14) | .753 |
| **Hospital discharge outcomes** |  |  |  |  |  |  |
| In-hospital mortality, n (%) | 3 (6.7) | 4 (4.1)<sup>b</sup> | 13 (9.1) | 8 (33.3)<sup>a</sup> | 28 (9.0) | < .001 |
| Home care referral, n (%), N = 282 | 1 (2.4) | 4 (4.3)<sup>b</sup> | 19 (14.6)<sup>a</sup> | 5 (31.1)<sup>a</sup> | 29 (10.3) | < .001 |
| Prolonged hospitalization (>13 days), n (%) | 16 (35.6) | 28 (28.6) | 52 (36.4) | 7 (29.2) | 103 (33.2) | .598 |
| **Post-discharge outcomes** |  |  |  |  |  |  |
| ED revisits or readmission, n (%), N = 282 | 9 (21.4)<sup>b</sup> | 37 (39.4) | 61 (46.9) | 9 (56.3) | 116 (41.1) | .017 |
| 90-day mortality, n (%), N = 310 | 4 (8.9)<sup>b</sup> | 14 (14.3)<sup>b</sup> | 36 (25.2) | 13 (54.2)<sup>a</sup> | 67 (21.6) | < .001 |
| 180-day mortality, n (%), N = 310 | 6 (13.3)<sup>b</sup> | 21 (21.4)<sup>b</sup> | 48 (33.6) | 15 (62.5)<sup>a</sup> | 90 (29.0) | < .001 |

**Abbreviations:** IVCF-20, Clinical-Functional Vulnerability Index-20; IQR, interquartile
range; CCI, Charlson Comorbidity Index; DM, Diabetes mellitus; MI, myocardial infarction;
TIA: transient ischemic attack; COPD, chronic obstructive pulmonary disease; CFS,
Clinical Frailty Scale; CKD, chronic kidney disease; ADL, activities of daily living; NEWS2,
National Early Warning Score 2; ED, Emergency Department. **Notes:** If the *P* - value was
significant, “a” indicates a frequency higher than expected and “b” indicates a frequency
lower than expected.

Clinical-functional frailty was associated with older age, female sex, and living with family members. The prevalence of frailty, as measured by the IVCF-20, was 53.9% (167 of 310; 95% CI, 48.1–59.5%). Using the CFS, excluding individuals with CFS 9 (terminal illness), the prevalence was 60.1% (170 of 283; 95% CI, 54.1–65.8%). Among frail individuals, mild frailty predominated according to the CFS (CFS 5 in 65.3% [111 of 170]).

When the CFS was used as a reference standard for identifying frailty in the ED, the AUC for the IVCF-20 was 0.93 (95% CI: 0.92–0.93; *P* < .001), indicating excellent sensitivity (81.3%) and specificity (91.1%) for frailty identification (Supplementary Figure S3). Regarding acute illness severity, higher frailty scores were significantly associated with higher median NEWS2 values. Similarly, comorbidity burden, as measured by the CCI, increased progressively and significantly with frailty status.

During the 6-month follow-up period, 90 older adults (29.0%) died. Of these, 67 deaths (21.6%) occurred within 90 days of hospital admission, and 28 deaths (9.0%) occurred during hospitalization. Frailty, as measured by the IVCF-20, was strongly associated with mortality across the three time points evaluated.

LOS ranged from 1 to 112 days. A total of 103 participants (33.2%) experienced prolonged LOS (>13 days, upper tertile), although no significant association with frailty was observed. Regarding institutionalization, three participants (1.0%) were admitted from long-term care facilities (LTCFs). One of these patients died during hospitalization, and the remaining two returned to their original LTCFs, but both died within 90 days of hospital admission. Among participants living with family members, only two (0.8%) were institutionalized within 3 months after discharge, whereas none of those living alone required institutionalization. IVCF-20 scores were not significantly associated with post-discharge institutionalization.

Among the 282 patients discharged alive, 29 (10.3%) were referred to home care services, and referral was significantly associated with higher IVCF-20 scores. At least one ED revisit was recorded for 116 participants (41.1%), of whom 77 (66.4%) required hospital readmission. The association between frailty and ED revisits or readmission was significant, with higher frailty levels associated with increased event frequency.

As an independent predictor for 180-day mortality, CFS yielded the higher AUC than IVCF-20 (AUC 0.75, 95% CI 0.69-0.82, *P* < .001 vs. AUC 0.65, 95% CI 0.59-0.72, *P* < .001) and this difference was statistically significant (DeLong test, *P* < .001).

In univariate analysis, the IVCF-20 score was strongly associated with 180-day mortality. Mortality rates by IVCF-20 category were as follows: 6 deaths (13.3%) for scores 0-6, 21 deaths (21.4%) for scores 7-14, 48 deaths (33.6%) for scores 15-29, and 15 deaths (62.5%) for scores 30-40. The unadjusted OR was 1.07 (95% CI, 1.04–1.11; *P* < .001), with an AUC of 0.65 (0.59–0.72).

Other variables significantly associated with 180-day mortality included metastatic cancer, dementia, delirium, chronic obstructive pulmonary disease, chronic liver disease, need help with bathing, impaired mobility, hospitalization within the previous 6 months, NEWS2 score, and CCI score (all *P* < .05). Unadjusted associations between baseline characteristics and 180-day mortality are presented in Table 2.

**Table 2.** Unadjusted analysis regarding 180-day mortality after admission to the Emergency Department.

| Variables | No<br>(n=220) | Yes<br>(n=90) | OR (95% CI) | P-value |
| --- | --- | --- | --- | --- |
| **Age** |  |  | 1.02 (0.99–1.06) | .101 |
| 60–74 | 140 (71.8) | 55 (28.2) |  |  |
| 75–84 | 61 (73.5) | 22 (26.5) |  |  |
| ≥85 | 19 (59.4) | 22 (26.5) |  |  |
| Male | 131 (59.5) | 49 (54.4) | 0.81 (0.49–1.33) | .409 |
| Non-white | 146 (66.4) | 54 (60.0) | 0.76 (0.46–1.26) | .288 |
| <8 years of education | 165 (75.0) | 68 (75.6) | 1.03 (0.58–1.82) | .918 |
| Live alone | 31 (14.1) | 7 (7.8) | 0.51 (0.22–1.21) | .124 |
| **IVCF-20 score** |  |  | 1.07 (1.04–1.11) | < .001 |
| IVCF-20 (categorical) |  |  |  |  |
| 7–14 vs. 0–6 | – | – | 1.78 (0.66–4.75) | .255 |
| 15–29 vs. 0–6 | – | – | 3.28 (1.30–8.30) | .012 |
| 30–40 vs. 0–6 | – | – | 10.83 (3.29–35.69) | < .001 |
| **CFS score** |  |  | 1.98 (1.65–2.37) | < .001 |
| 1–3 | 47 (88.7) | 6 (11.3) |  |  |
| 4 | 50 (84.7) | 9 (15.3) |  |  |
| 5–6 | 111 (78.2) | 31 (21.8) |  |  |
| 7–9 | 12 (21.4) | 44 (78.6) |  |  |
| **CCI score** |  |  | 1.37 (1.23–1.52) | < .001 |
| 0–5 | 105 (86.8) | 16 (13.2) |  |  |
| 6 | 36 (67.9) | 17 (32.1) |  |  |
| 7–8 | 44 (65.7) | 23 (34.3) |  |  |
| ≥9 | 35 (50.7) | 34 (49.3) |  |  |
| DM with end-organ damage | 38 (17.3) | 10 (11.1) | 0.60 (0.28–1.26) | .173 |
| History of myocardial infarction | 48 (21.8) | 17 (18.9) | 0.83 (0.45–1.55) | .565 |
| Heart failure | 80 (36.4) | 28 (31.1) | 0.79 (0.47–1.33) | .378 |
| Stroke/ transient ischemic attack | 38 (17.3) | 14 (15.6) | 0.88 (0.45–1.72) | .713 |
| Metastatic cancer | 13 (5.9) | 33 (36.7) | 9.22 (4.55–18.7) | < .001 |
| Dementia | 16 (7.3) | 14 (15.6) | 2.35 (1.09–5.04) | .025 |
| Delirium | 9 (4.1) | 15 (16.7) | 4.69 (1.97–11.2) | < .001 |
| Chronic obstructive pulmonary disease | 30 (13.6) | 21 (23.3) | 1.93 (1.03–3.59) | .037 |
| Chronic liver disease | 13 (5.9) | 14 (15.6) | 2.93 (1.32–6.52) | .006 |
| CKD (moderate to severe) | 54 (24.5) | 27 (30.0) | 1.32 (0.76–2.27) | .321 |
| Need help with bathing | 27 (12.3) | 30 (33.3) | 3.57 (1.97–6.48) | < .001 |
| Impaired mobility (ADL limitation) | 109 (49.5) | 67 (74.4) | 2.97 (1.72–5.10) | < .001 |
| Polypharmacy (≥5 medications) | 143 (65.0) | 63 (70.0) | 1.26 (0.74–2.13) | .397 |
| Hospitalization in previous 6 months | 105 (47.7) | 64 (71.1) | 2.70 (1.59–4.57) | < .001 |
| ≥2 falls in the past year | 56 (25.5) | 15 (16.7) | 0.59 (0.31–1.10) | .095 |
| **NEWS2 score** |  |  | 1.26 (1.14–1.39) | < .001 |
| 0–1 | 91 (83.5) | 18 (16.5) |  |  |
| 2 | 35 (76.1) | 11 (23.9) |  |  |
| 3–5 | 63 (66.3) | 32 (33.7) |  |  |
| ≥6 | 31 (51.7) | 29 (48.3) |  |  |
| Length of stay (continuous) | – | – | 1.01 (0.99–1.03) | .143 |

**Abbreviations:** OR, odds ratio; CI, confidence interval; IVCF-20, Clinical-Functional
Vulnerability Index-20; CFS, Clinical Frailty Scale; CCI, Charlson Comorbidity Index; DM, Diabetes mellitus; CKD, chronic kidney disease; ADL, activities of daily living; NEWS2, National Early Warning Score 2. **Notes:** *P* - values and OR are based on original values and not on stratified groups. Analyses reported both with IVCF-20 treated as a continuous and as a categorical variable, respectively.

In the multivariate analysis, the IVCF-20 treated as a continuous variable, was significantly associated with 180-day mortality (aOR = 1.06; 95% CI, 1.02–1.10; *P* = .002). In a second model, the IVCF-20 was included as an ordinal categorical variable to facilitate interpretation of the mortality gradient across increasing frailty levels. Using robust older adults (IVCF-20 0–6) as the reference group, a progressive increase in the odds of 180-day mortality was observed with higher frailty, although statistical significance was reached only in the most severely frail category (IVCF-20 30–40).

To better estimate the excess risk associated with severe frailty, additional analysis compared the severe frailty group (IVCF-20 30–40) with each of the other categories, adjusting the reference category for each comparison. Severely frail older adults had 8.4 times higher odds of death compared with robust individuals (OR = 8.37; 95% CI, 2.20–31.81), 5.6 times higher odds compared with pre-frail participants (OR = 5.64; 95% CI, 1.92–16.62), and 3.7 times higher odds compared with those with mild-to-moderate frailty (OR = 3.69; 95% CI, 1.37– 9.97). Kaplan-Meier survival curves confirmed the progressive increase in mortality risk with higher IVCF-20 scores (Figure 1), with statistical significance demonstrated by the log-rank test (χ²(3) = 32.1; *P* < .001).

**Figure 1.**
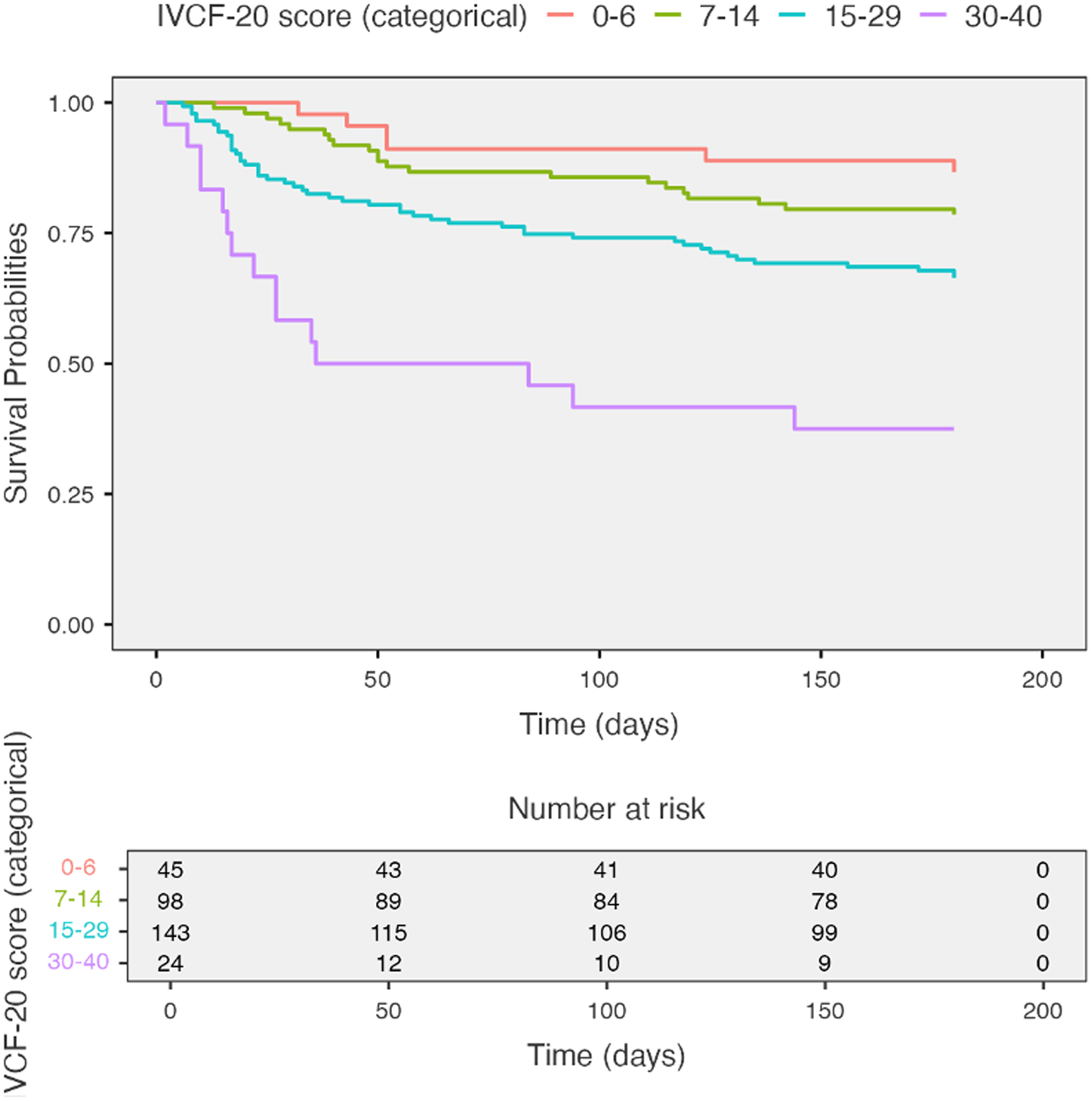
Kaplan–Meier survival curves according to frailty status. Kaplan–Meier survival estimates for 180-day mortality by IVCF-20 categories: robust (0–6), pre-frail (7–14), mild-to-moderate frailty (15–29), and severe frailty (30–40). Survival probability decreased progressively with increasing frailty, with a statistically significant difference across groups (log-rank test, χ²(3) = 32.1; *P* < .001).

The CFS was also significantly associated with 180-day mortality. Mortality rates were 6 deaths (11.3%) for CFS 1-3, 9 deaths (15.3%) for CFS 4; 31 deaths (21.8%) for CFS 5-6; and 44 deaths (78.6%) for CFS 7-9. The unadjusted OR was 1.98 (95% CI, 1.65–2.37; *P* < .001), with an AUC of 0.75 (0.69–0.82). In multivariate analysis, the CFS remained significantly associated with 180-day mortality (aOR = 1.66; (95% CI, 1.34–2.04; *P* < .001) (Table 3).

**Table 3.**
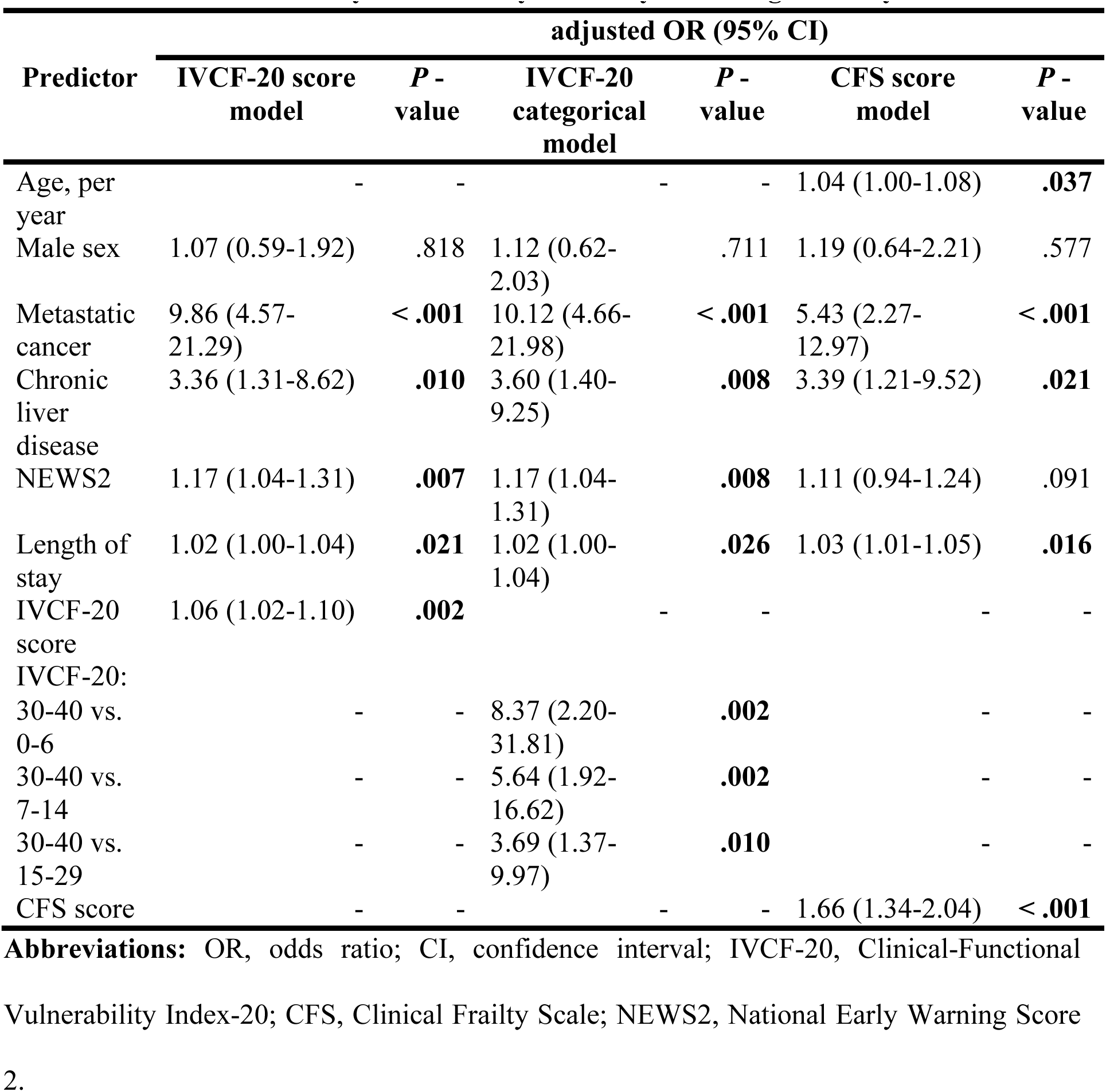
Multivariable analysis of 180-day mortality according to frailty instruments.

| Predictor | adjusted OR (95% CI) |  |  |  |  |  |
| --- | --- | --- | --- | --- | --- | --- |
|  | IVCF-20 score model | *P* - value | IVCF-20 categorical model | *P* - value | CFS score model | *P* - value |
| Age, per year | - | - | - | - | 1.04 (1.00-1.08) | **.037** |
| Male sex | 1.07 (0.59-1.92) | .818 | 1.12 (0.62-2.03) | .711 | 1.19 (0.64-2.21) | .577 |
| Metastatic cancer | 9.86 (4.57-21.29) | **< .001** | 10.12 (4.66-21.98) | **< .001** | 5.43 (2.27-12.97) | **< .001** |
| Chronic liver disease | 3.36 (1.31-8.62) | **.010** | 3.60 (1.40-9.25) | **.008** | 3.39 (1.21-9.52) | **.021** |
| NEWS2 | 1.17 (1.04-1.31) | **.007** | 1.17 (1.04-1.31) | **.008** | 1.11 (0.94-1.24) | .091 |
| Length of stay | 1.02 (1.00-1.04) | **.021** | 1.02 (1.00-1.04) | **.026** | 1.03 (1.01-1.05) | **.016** |
| IVCF-20 score | 1.06 (1.02-1.10) | **.002** | - | - | - | - |
| IVCF-20: 30-40 vs. 0-6 | - | - | 8.37 (2.20-31.81) | **.002** | - | - |
| 30-40 vs. 7-14 | - | - | 5.64 (1.92-16.62) | **.002** | - | - |
| 30-40 vs. 15-29 | - | - | 3.69 (1.37-9.97) | **.010** | - | - |
| CFS score | - | - | - | - | 1.66 (1.34-2.04) | **< .001** |

**Abbreviations:** OR, odds ratio; CI, confidence interval; IVCF-20, Clinical-Functional
Vulnerability Index-20; CFS, Clinical Frailty Scale; NEWS2, National Early Warning Score

In the comparative AUC analysis, the CFS model demonstrated better predictive performance than the model using the IVCF-20 as a continuous variable (DeLong test, *P* = .037). No significant difference was observed when comparing the CFS model with the model using IVCF-20 as a categorical variable (DeLong test *P* = .197) (Supplement Figure S4).

### Secondary Outcomes

The AUC analysis was used to compare the ability of the IVCF-20 and the CFS to independently predict adverse outcomes, including in-hospital mortality, 90-day mortality, home care referral, and ED revisit or readmission within 3 months after discharge. The CFS consistently demonstrated higher AUC values than the IVCF-20 for all outcomes assessed. This difference was statistically significant for mortality (DeLong test, *P* < .001 for 90-day mortality; *P* = .009 for in-hospital mortality), but not significant for home care referral (DeLong test, *P* = .076) or for ED revisits/readmissions (DeLong test, *P* = .289) (Table 4 and Supplementary figures from S5 to S8 for AUCs).

**Table 4.**
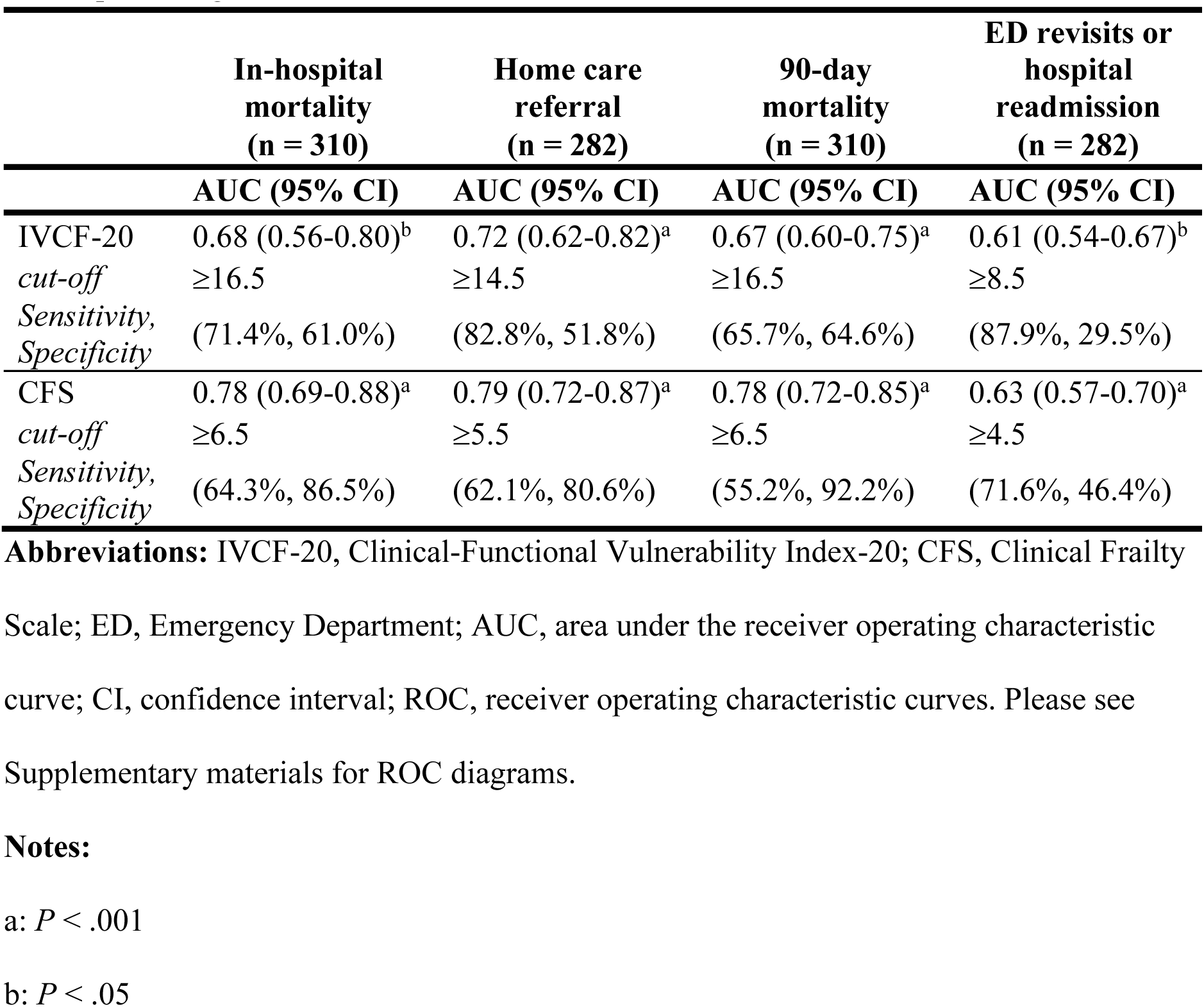
The Area Under the Receiver Operating Characteristic Curves for IVCF-20 and CFS for the predicting adverse outcomes.

## Discussion

In this study, we demonstrated that the IVCF-20, a rapid and multidimensional frailty screening tool, was able to predict mortality and other adverse outcomes in older adults admitted to the ED. The predictive ability of the IVCF-20 was comparable to that of other rapid frailty instruments ^18,32^. As a continuous variable, it behaved as a linear and independent predictor of 180-day mortality after hospital admission, with a clear gradient of increasing mortality across higher frailty scores. As a categorical variable, we proposed a new subcategory among the most frail (severe frailty, IVCF-20 30–40), which allowed quantification of how this group differed from others, showing substantially higher odds of death within 6 months. The IVCF-20 score, combined with acute deterioration criteria measured by NEWS2 and presence of severe comorbidities, showed good accuracy in predicting 180-day mortality. Previous studies have shown that combining baseline clinical-functional vulnerability with acute deterioration and severe comorbidities may reduce prognostic uncertainty ^33–36^.

The prevalence of frailty among hospitalized older adults can be up to 4 times higher than in community-dwelling older adults. In our study, the prevalence of frailty in the ED was 53.9% by IVCF-20 and 60.1% by CFS, consistent with previous studies ^11,37^. In the emergency setting, pooled prevalence estimates from systematic reviews and meta-analyses range between 20% to 91% ^38^. This wide variability reflects the large number of frailty instruments used and differences in age distribution among study populations. Studies including only older adults >70 years reported higher prevalence rates. In our study, most participants were younger than 75 years, but frailty was significantly more prevalent in those aged ³ 75 years (72.8% vs. 50.3%).

In our sample, 180-day mortality reached 62.5% among older adults with IVCF-20 scores between 30 and 40. Identifying baseline frailty through preadmission IVCF-20 scores at the time of ED admission can assist hospital teams in defining care plans and avoiding invasive, non-beneficial, or potentially harmful treatments for severely frail older adults, who may instead benefit from palliative approaches.

We observed mortality rates higher than previously reported ^39–42^. This likely reflects the case mix at the HU-UFMG, where patients with severe and advanced chronic diseases predominate. Advanced cancer and chronic liver disease were strong predictors of 180-day mortality and remained in all final models.

Previous research has emphasized that, in the ED, a frailty instrument must be accurate, easy and quick to apply, multidimensional, independent of physical performance measures or technology, reproducible, easy to interpret, and suitable for longitudinal monitoring ^18,19,43^. The IVCF-20 meets these criteria: it is valid, easy to administer, highly correlated with the CGA, requires minimal technology and training, and does not rely on clinical judgment. However, the IVCF-20 had not previously been evaluated in ED or for outcome prediction. It is widely used in primary health care across several Brazilian cities ^44–47^ and is included in the National Ministry of Health’s Manual of Multidimensional Assessment of Older Adults for Primary Care ^48^, which could allow the identification of baseline frailty as assessed in primary care prior to ED admission. Using the same instrument across care settings could facilitate longitudinal monitoring. Furthermore, with the support of integrated electronic health records, information from primary care can be shared with acute care settings, potentially enabling better care planning and management for frail older adults. These findings support the feasibility and potential clinical utility of the IVCF-20 for frailty screening in time-limited settings, such as Emergency Departments.

In high-demand ED settings, the main barriers to implementing frailty instruments are limited time, lack of training, and a reliance on clinical judgment ^18^. ELLIOTT et al. ^19^ assessed the feasibility of four frailty instruments in the ED, including the CFS, and found that some professionals preferred relying solely on clinical judgment. However, isolated clinical judgment was less accurate than when combined with frailty instruments. In our study, the predictive ability of the CFS was superior to that of the IVCF-20. Nevertheless, this difference was not significant when excluding CFS item 9. This highlights the influence of clinical judgment in enhancing the predictive ability of the CFS and its greater subjectivity in classifying the terminal illness category.

Further research is needed to validate the IVCF-20’s predictive ability for outcomes and to support its application across different points of the healthcare system.

### Limitations

This study has some limitations. Due to limited human resources during the first phase of data collection and restrictions imposed by the COVID-19 pandemic, participant inclusion was limited to weekdays between 8:00 a.m. and 5:00 p.m., which reduced the sample size and may have introduced selection bias. Patients discharged on the same day of ED admission - likely with less severe acute conditions - were not included, limiting the generalizability of results to this subgroup. The study sample consisted predominantly of patients with a high burden of severe chronic diseases (e.g., advanced cancer, liver disease, and heart disease), as HU-UFMG is a quaternary care center. Therefore, caution is warranted when extrapolating these findings to patients with lower clinical complexity.

Nevertheless, this study has important strengths. There were no missing data for mortality outcomes, and we used validated and reliable frailty instruments to ensure measurement consistency.

## Conclusion

This study confirmed the high prevalence of frailty among older adults admitted to the Emergency Department and demonstrated that frailty, as measured by the IVCF-20, predicts 180-day mortality, home care referral, and Emergency Department revisits or hospital readmission within 3 months after discharge. The IVCF-20 is a multidimensional, rapid, and easy-to-use frailty screening tool that does not rely on clinical judgment, making it feasible for implementation in emergency settings. Its integrated use across primary and acute care could enhance continuity of frailty assessment and promote coordinated care planning for frail older adults. Further studies are needed to validate these findings in populations with different characteristics.

## Data Availability

All data produced in the present study are available upon reasonable request to the authors

## Acknowledgments

We thank all participants of this study; the CHANGE Study for the support in this study, UH-UFMG for its logistics and infrastructure support, and the financial support entities: Conselho Nacional de Desenvolvimento e Pesquisa (CNPq), Coordenação de Aperfeiçoamento de Pessoal de Nível Superior (CAPES), Fundação de Amparo à Pesquisa do Estado de Minas Gerais (FAPEMIG).

## Author contributions

Tatiana C. E. Pinheiro: study concept and design, acquisition of data, data analysis and interpretation, and manuscript preparation. Marco A. F. Angelo: data analysis, data interpretation. Márlon J. R. Aliberti: study concept and design, manuscript revision. Carla J. Machado: data analysis, manuscript revision. Vicente E. Carvalho: acquisition of data, manuscript preparation. Nádia E. Ramos: acquisition of data, manuscript preparation. Camila O. Alcântara: acquisition of data, manuscript preparation. Fabiano M. Pereira: acquisition of data, manuscript preparation. Ana P. A. F. Peixoto: acquisition of data, manuscript preparation. Fátima S. Machado: acquisition of data, manuscript preparation. Bernardo M. Viana: manuscript revision. Edgar N. Moraes: manuscript revision. Maria A. C. Bicalho: study concept and design, data interpretation, and manuscript preparation and revision.

