## Supplement Figures for "The Clinical-Functional Vulnerability Index-20 (IVCF-20) Predicts Adverse Outcomes in Older Adults Admitted through the Emergency Department"

### Supplementary materials

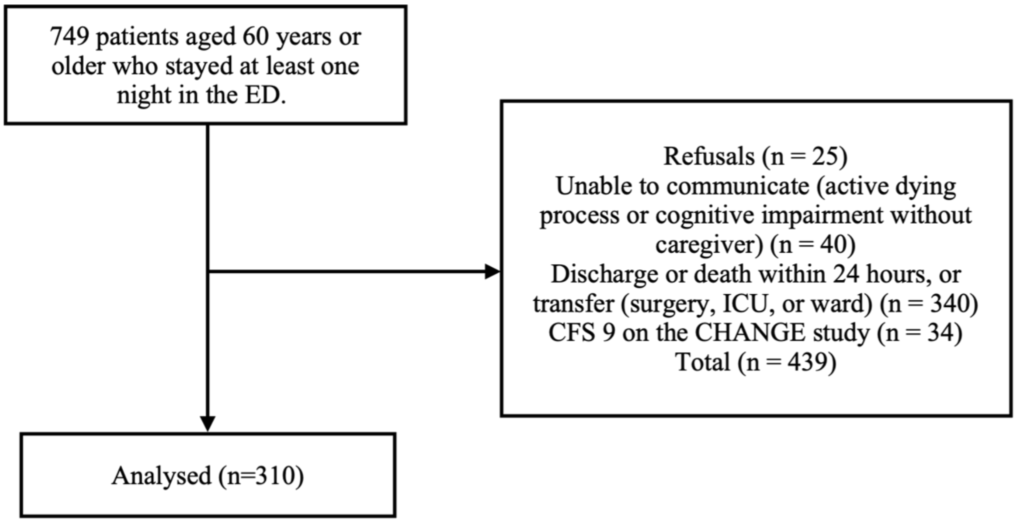

Note: Abbreviations: ED, Emergency Department; ICU, intensive care unit; CFS, Clinical Frailty Scale; CHANGE, Creating a Hospital Assessment Network in Geriatrics.

**Supplementary Figure S1 -** Flow diagram of participant selection and follow-up.

| CLINICAL-FUNCTIONAL VULNERABILITY INDEX – 20 (CFVI-20)<br>www.ivcf20.org | | | |
|---|---|---|---|
| **AGE** | | **1.** How old are you? | ( ) 60-74 years *(0)* |
| | | | ( ) 75-84 years *(1)* |
| | | | ( ) ≥ 85 years *(3)* |
| **HEALTH SELF-PERCEPTION** | | **2.** In general, compared with others your age, how would you rate your health? | ( ) Very good or Good *(0)* |
| | | | ( ) Fair or poor *(1)* |
| **INSTRUMENTAL ADL**<br>*The maximum score for this item is 4 points, even if the older person answered yes to all questions 3, 4 and 5.* | | **3.** Because of your health or physical condition, have you stopped shopping?<br>( ) Yes (4) ( ) No or you no longer shop for reasons other than health (0)<br>**4.** Because of your health or physical condition, have you stopped managing your money, spending, or paying your household bills?<br>( ) Yes (4) ( ) No or you no longer manage your money for reasons other than health (0)<br>**5.** Because of your health or physical condition, have you stopped doing small household chores, such as washing dishes, tidying the house, or doing light cleaning?<br>( ) Yes (4)<br>( ) No or you no longer do small household chores for reasons other than health (0) | |
| **Basic ADL** | | **6.** Because of your health or physical condition, have you stopped bathing yourself? ( ) Yes (6) ( ) No (0) | |
| **COGNITION** | | 7. Has a family member or friend told you that you are becoming forgetful?<br>( ) Yes (1) ( ) No (0) | |
| | | **8.** Has this forgetfulness gotten worse in recent months? ( ) Yes (1) ( ) No (0) | |
| | | **9.** Is this forgetfulness preventing you from carrying out any daily activities?<br>( ) Yes (2) ( ) No (0) | |
| **HUMOR** | | **10.** In the last month, have you felt discouraged, sad, or hopeless?<br>( ) Yes (2) ( ) No (0) | |
| | | **11.** In the last month, have you lost interest or pleasure in activities you usually enjoy? ( ) Yes (2) ( ) No (0) | |
| **MOBILITY** | **Reach, grip and pinch** | **12.** Are you unable to raise your arms above shoulder level? ( ) Yes (1) ( ) No (0) | |
| | | **13.** Are you unable to handle or hold small objects? ( ) Yes (1) ( ) No (0) | |
| | **Aerobic and/or muscular capacity**<br>*The maximum score for this item is 2 points, even if the individual answered yes to all four questions.* | **14.** Do you have any of the three conditions listed below? ( ) Yes (2) ( ) No (0)<br>• Unintentional weight loss of 4.5 kg or 5% of body weight in the last year or 6 kg in the last 6 months or 3 kg in the last month ( )<br>• Body Mass Index (BMI) less than 22 kg/m2 ( )<br>• Calf circumference (perimeter) < 31 cm ( )<br>• Time spent on the gait speed test (4 m) > 5 sec ( ) | |
| | **Gait** | **15.** Do you have difficulty walking that could prevent you from carrying out some daily activities? ( ) Yes (2) ( ) No (0) | |
| | | **16.** Have you had two or more falls in the last year? ( )Yes (2) ( )No (0) | |
| | **Sphincter Continence** | **17.** Do you ever accidentally leak urine or feces? ( ) Yes (2) ( ) No (0) | |
| **COMUNICATION** | **Vision** | **18.** Do you have vision problems that interfere with daily activities?<br>Glasses or contact lenses are permitted. ( ) Yes (2) ( ) No (0) | |
| | **Hearing** | **19.** Do you have hearing problems that interfere with daily activities?<br>Hearing aids are permitted. ( ) Yes (2) ( ) No (0) | |
| **MULTIPLE COMORBIDITY**<br>*The maximum score for this question is 4 points, even if the older adult answered yes to all three questions.* | | **20.** Do you have any of the three conditions listed below? ( ) Yes (4) ( ) No (0)<br>• Five or more chronic diseases.<br>• Regular use of five or more different medications daily.<br>• Recent hospitalization within the last 6 months. | |
| | | | **Final score** |

**Supplementary Figure S2** – Clinical-Functional Vulnerability Index-20, English version.

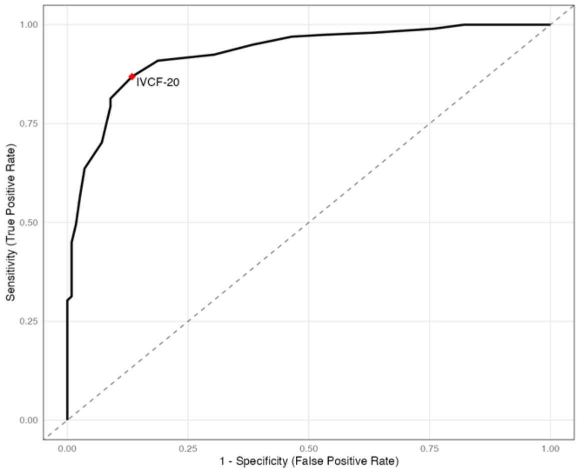

| | AUC (95% CI) | *P* - value | Cut point (Sensitivity, Specificity) |
|---|---|---|---|
| **IVCF-20** | 0.93 (0.92-0.93) | < .001 | ≥ 13.5 (81.3%, 91.1%) |

Note: Abbreviations: AUC, area under the receiver operating characteristic curves; IVCF-20, Clinical-Functional Vulnerability Index.

**Supplementary Figure S3 -** The AUCs for IVCF-20 against the reference Clinical Frailty Scale in diagnosis of frailty.

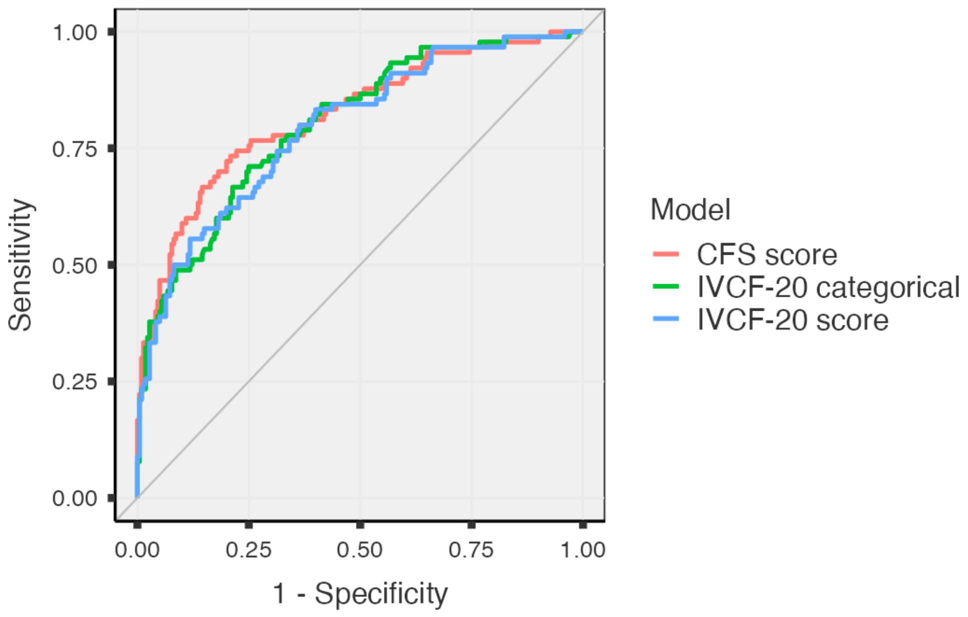

| **Models** | **AUC (95% CI)** |
|---|---|
| **CFS score** | 0.82 (0.77-0.87) |
| **IVCF-20 categorical** | 0.80 (0.75-0.86) |
| **IVCF-20 score** | 0.79 (0.74-0.85) |

Note: Abbreviations: AUC, area under the receiver operating characteristic curves; CFS, Clinical Frailty Scale; IVCF-20, Clinical-Functional Vulnerability Index.

**Supplementary Figure S4 -** AUCs comparing the predictive performance of the CFS and the IVCF-20 models for 180-day mortality.

#### Secondary outcomes

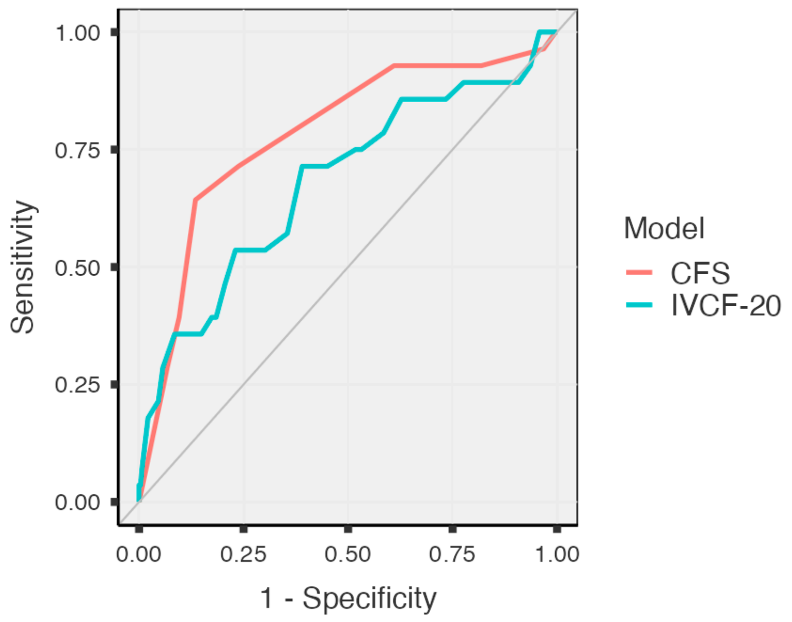

**Supplementary Figure S5 -** AUCs for Clinical Frailty Scale (CFS) and Clinical-Functional Vulnerability Index-20 (IVCF-20) predicting in-hospital mortality.

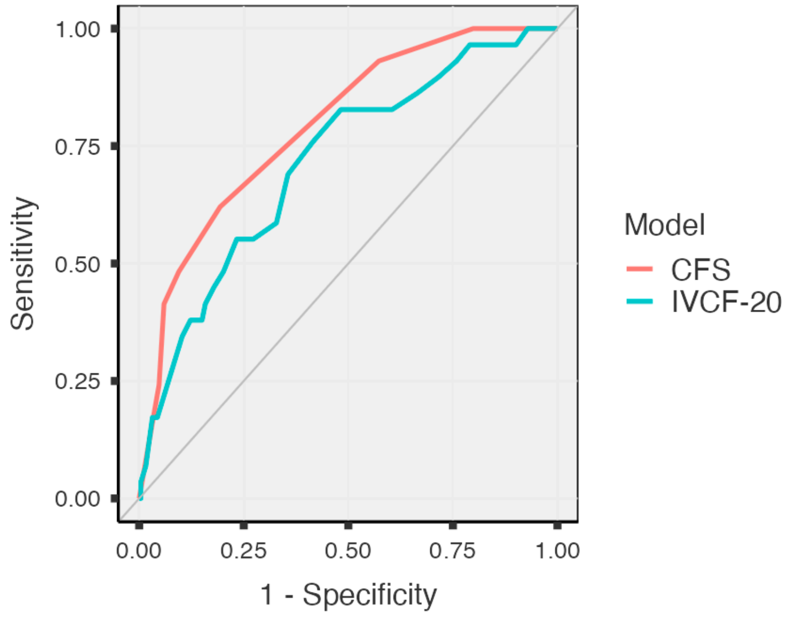

**Supplementary Figure S6 -** AUCs for Clinical Frailty Scale (CFS) and Clinical-Functional Vulnerability Index-20 (IVCF-20) predicting home care referral.

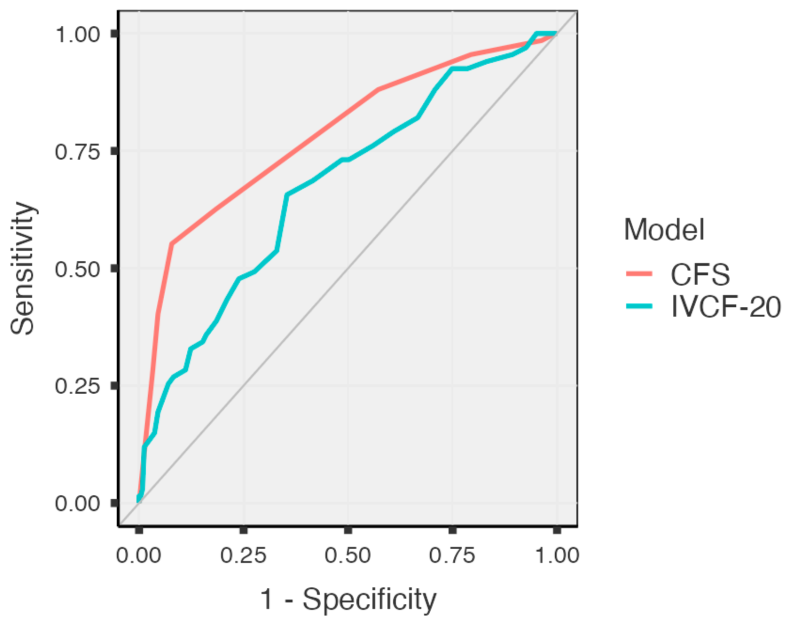

**Supplementary Figure S7 -** AUCs for Clinical Frailty Scale (CFS) and Clinical-Functional Vulnerability Index-20 (IVCF-20) predicting 90-day mortality.

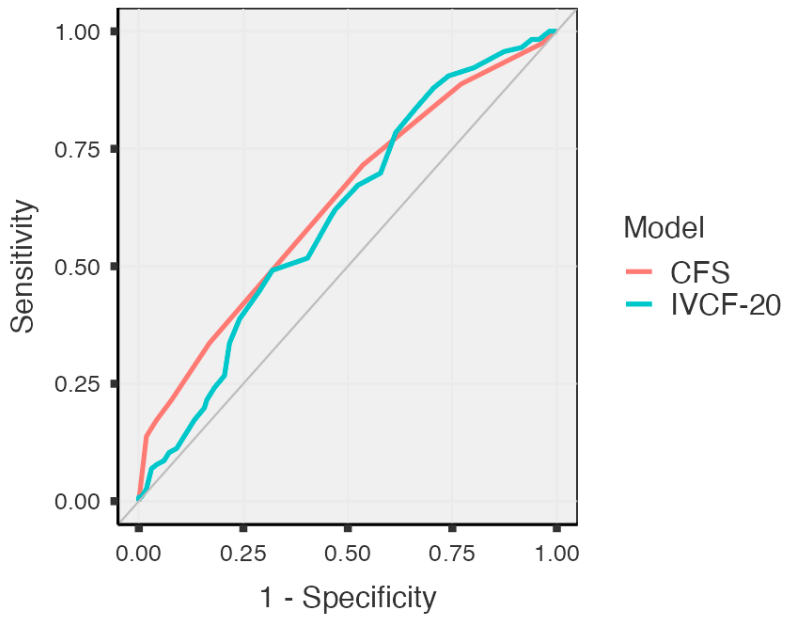

**Supplementary Figure S8 -** AUCs for Clinical Frailty Scale (CFS) and Clinical-Functional Vulnerability Index-20 (IVCF-20) in predicting ED revisits or hospital readmission.
